# Segmental infralesional lower motor neuron abnormalities in patients with sub-acute traumatic spinal cord injury

**DOI:** 10.1101/2023.02.18.23286121

**Authors:** Michael J. Berger, Adenike A. Adewuyi, Christopher Doherty, Amy K. Hanlan, Cynthia Morin, Russ O’Connor, Radhika Sharma, Shannon Sproule, Kevin N. Swong, Harvey Wu, Colin K. Franz, Erin Brown

## Abstract

The health of the infralesional lower motor neuron (LMN) has received little attention in individuals with traumatic cervical spinal cord injuries (SCI). Infralesional LMN health is clinically relevant in the context of nerve transfer surgery to restore critical upper limb functions, as those demonstrating LMN damage below the neurological level of injury may experience irreversible sequelae of denervation (e.g., atrophy, fibrosis) without timely intervention. In this two-centre retrospective cohort study, we examined the health of the infralesional LMN in individuals with traumatic cervical SCI, using data derived from the clinical electrodiagnostic examination performed early after SCI. We assessed 66 limbs in 42 individuals with traumatic cervical SCI (40 males, mean age = 43.6±17.2, mean duration from injury = 3.3±1.5 months, 25 with motor complete injuries). Analysis was stratified by injury level as 1) C4 and above, 2) C5 and 3) C6-7. EMG performed on representative muscles from C5-6, C6-7, C7-8 and C8-T1, were included in analysis. LMN abnormality was dichotomized as present (abnormal spontaneous activity) or absent. Data were pooled for the most caudal infralesional segment (C8-T1). Overall, a high frequency of denervation potentials was seen in all infralesional segments for all injury levels. The pooled frequency of denervation potentials at C8-T1 was 74.6% of limbs tested. There was also evidence of denervation potentials at the rostral border of the neurological level of injury, as high as 64.3% of C5-6 muscles for C5 injuries. These data support a high prevalence of infralesional LMN abnormality following SCI, which has implications to candidacy, timing of the intervention, donor nerve options and motor prognosis following SCI.

## Introduction

Investigations on the origins of muscle weakness and paralysis in traumatic spinal cord injury (SCI) typically emphasize the damaged cortical spinal tracts, at the spinal lesion epicenter^1^ Less weight has been placed on understanding the impact of the spinal lesion on the lower motor neuron (LMN).

It is well-established that SCI has significant downstream effects on skeletal muscle, including muscle fibre atrophy, fibre-type switching and changes in metabolism, with these changes being attributed to the chronic absence of supraspinal input.^2^ Yet, there is a growing understanding that the LMN may be more significantly and directly altered in traumatic SCI. Understanding the impact of SCI on the peripheral neuromuscular system is critically important, especially in the context of novel interventions to address motor function. Of particular recent interest is nerve transfer surgery to restore vital hand and upper limb functions in those with cervical SCI.^3,4^ The success of nerve transfer surgery - re-routing a functionally redundant donor nerve from above the neurological level of injury, to a paralyzed recipient muscle group below the injury – is based on the premise that the infralesional LMN (below the neurological level of injury) is anatomically and physiologically unaltered.^5^ To date, only a few small studies have investigated the health of the LMN in the infralesional zone and they raise concerns that LMN status is frequently altered after SCI.^6-8^

The clinical electrodiagnostic examination can be used to localize and determine the severity of LMN lesions in SCI, yet individuals with cervical SCI are not routinely referred for LMN assessment. Other methods for LMN evaluation in SCI, including with surface electrical stimulation, have also been proposed.^9^ Studies investigating LMN health caudal to the injury level have demonstrated evidence of muscle membrane instability (presence of fibrillations and positive sharp waves).^10,11^ Other investigations into LMN health have shown reduced motor neuron numbers at^12^ and caudal to the lesion site.^13,14^ The majority of these investigations have occurred in individuals with longstanding (i.e., chronic) injuries, and in muscle groups that are not clinically relevant to nerve transfer surgery. Understanding LMN health soon after injury, and in clinically relevant muscles, is important in the context of nerve transfer. LMN degeneration results in irreversible motor endplate degradation and muscle fibrosis without early intervention.^15,16^ If LMN damage is not identified, the critical time window for intervention with nerve transfer may be missed.

In this study, we retrospectively examined the frequency and longitudinal extent of infralesional LMN abnormality in cervical segments, in individuals with sub-acute cervical SCI who were undergoing evaluation for nerve transfer surgery candidacy (within 6 months of injury). We hypothesized that LMN abnormality would occur with a high frequency and be observed caudal to the injury level (i.e., outside of the lesional zone).

## Methods

We employed a two-centre retrospective cohort design. Data were collected at two tertiary-hospital interdisciplinary upper limb programs for individuals with SCI, between 2018-2022. Ethical approval for retrospective data analysis was obtained from the local institutional ethical review boards. All individuals included in the analysis had cervical-level SCI and American Spinal Injuries Association Impairment Scale (AIS) A-D. Neurological level of injury was determined according to the International Standards for Neurological Classification of Injury^17^, which was obtained from the examination performed by their attending physiatrist upon admission to inpatient rehabilitation. Subjects were included if they underwent needle electromyography (EMG) examination in at least one upper limb, within six months of their injury. Six months was selected as this is understood to be the “critical window” for attempt at reinnervation through nerve transfer, to avoid the irreversible effects of chronic denervation.^18^ Each limb was analyzed as a single event (those who had both limbs tested counted as two events).

In this study, we focused on reporting the needle EMG findings, rather than nerve conduction studies (NCS). While NCS provides more quantitative information about LMN health,^19^ there are several limitations to upper limb NCS in the context of SCI and nerve transfer: readily available NCS with widely accepted normative data (e.g., median nerve stimulation with recording over the abductor pollicis brevis; ulnar nerve stimulation with recording over the first dorsal interosseous or abductor digit minimi) only evaluate the C8-T1 spinal segments and median and ulnar innervated muscles are not typically the recipients of nerve transfer surgery in SCI and 3) median and ulnar nerves are prone to focal entrapment, unrelated to the pathophysiology of SCI. However, we secondarily determined the relationship between the presence of LMN abnormality as defined by the needle EMG findings and the baseline-to-peak amplitude of the compound muscle action potential (CMAP) of the median motor study. This relationship was only examined for the C8-T1 segments, as the median motor study to the abductor pollicis brevis muscle derives its innervation from these segments. The cutoff for normal was a CMAP baseline-to-peak amplitude of 4.5 mV.^20^

LMN abnormality was determined dichotomously as the presence versus absence of muscle membrane instability on the needle EMG examination, as evidenced by fibrillation potentials and/or positive sharp waves. Even if a subject was able to volitionally activate a muscle supplied by an infralesional segment and there was evidence of neurogenic changes (e.g., polyphasic motor unit potentials, reduced recruitment), a muscle was only deemed to be “abnormal” in the presence of abnormal spontaneous activity. This was done to reduce the subjectivity associated with the needle examination.

Determining the involved spinal segments corresponding to the muscles sampled, was based on the neuroanatomic principle that no muscle is supplied by single spinal root, therefore data were classified according to overlapping spinal levels, as shown in Table 1. We also sought to determine the frequency of denervation potentials observed at contiguous segments caudal to the clinical neurological of injury up until the T1.

**Table 1.**
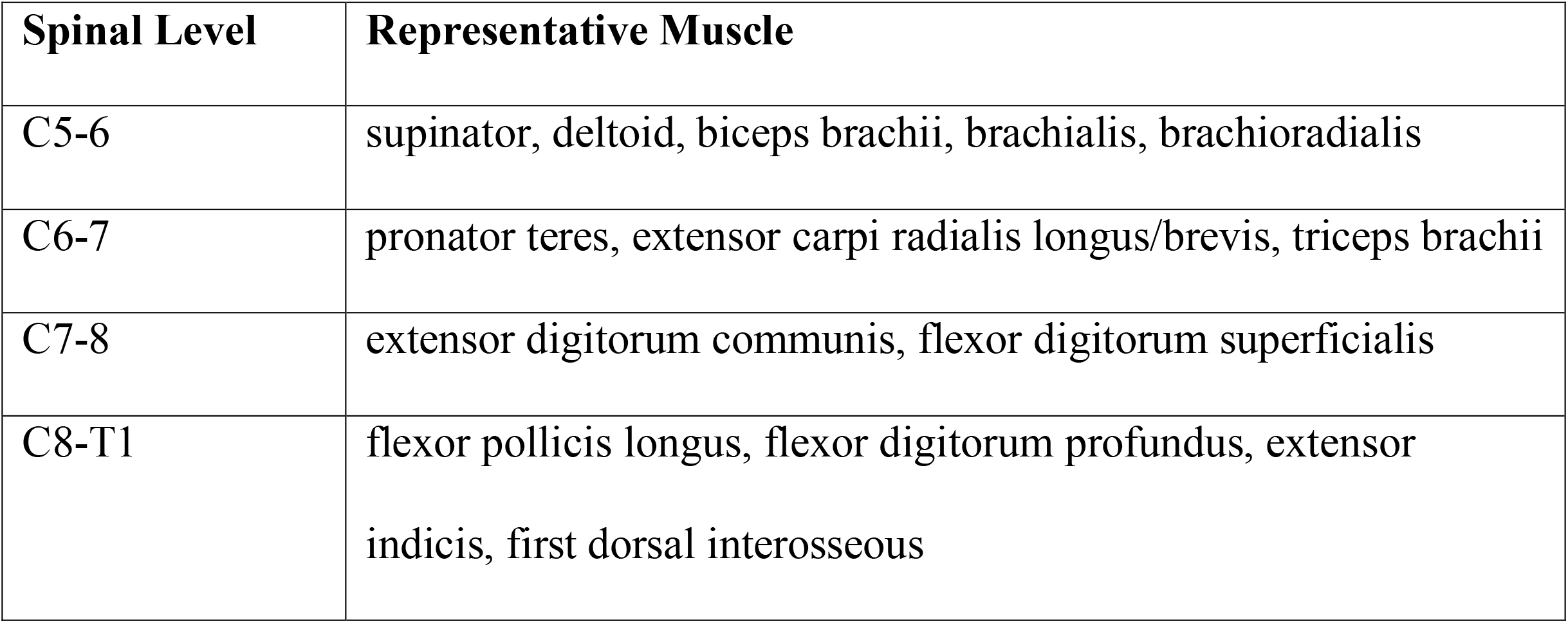
Classification of spinal level according to representative muscle tested.

| **Spinal Level** | **Representative Muscle** |
| --- | --- |
| C5-6 | supinator, deltoid, biceps brachii, brachialis, brachioradialis |
| C6-7 | pronator teres, extensor carpi radialis longus/brevis, triceps brachii |
| C7-8 | extensor digitorum communis, flexor digitorum superficialis |
| C8-T1 | flexor pollicis longus, flexor digitorum profundus, extensor indicis, first dorsal interosseous |

### Statistics

All descriptive data were presented as means and standard deviations. Frequency distributions were used to characterize the presence of LMN denervation. To compare the findings of the needle EMG and CMAP for the C8-T1 levels, we grouped data into a 2×2 matrix and used McNemar’s test to determine the concordance between EMG and CMAP data.

## Results

Demographic information is presented in Table 2. The majority of participants were male and had AIS motor complete injury (AIS A or B). A total of 66 limbs were analyzed in 42 individuals. The distribution of injury levels was C4 and above: 25 (59.5%), C5: 9 (21.4%), C6: 7 (16.6%) and C7: 1 (2.3%). Due to heterogeneity in injury level, sub-cohort analysis was performed for those with: 1) C4 level injuries and above, 2) C5 level injuries and 3) C6-7 level injuries. Note that C6 and C7 injury levels were combined in a single bin, as only a single individual with C7 injury level was included in the analysis. Data were only pooled for the C8-T1 spinal segments, which were caudal to the neurological level of injury for all individuals included in the study. The frequency distributions of denervation potentials observed for each injury level are depicted in Figure 1. Not all participants had all relevant segments evaluated and the number of observations for each spinal segment are listed in parentheses in Figure 1. For those with C4 level injuries and above, the frequencies of observed denervation potentials were 78.3%, 75.8%, 67.9% and 68.6% for the C5-6, C6-7, C7-8 and C8-T1 spinal levels, respectively. For those with C5 level injuries, the frequencies of observed denervation potential were 64.3%, 83.8%, 80.0% and 80.0% for the C5-6, C6-7, C7-8 and C8-T1 spinal levels, respectively. For those with C6-7 level injuries, the frequencies of observed denervation potential were 25.0%, 83.3%, 100% and 86.7% for the C5-6, C6-7, C7-8 and C8-T1 spinal levels, respectively. For pooled data at the C8-T1 levels, the frequency of LMN abnormality was observed to be 74.6%.

**Table 2.** Demographic information.

|  |  |
| --- | --- |
| Sex | 40 male, 2 female |
| Age (mean±standard deviation) | 43.6±17.2 |
| Months from injury (mean±standard deviation; range) | 3.3±1.5; 0.75-6.5 |
| *AIS* |  |
| A | 18 |
| B | 7 |
| C | 8 |
| D | 9 |

AIS: American Spinal Injuries Association Impairment Scale

**Figure 1.**
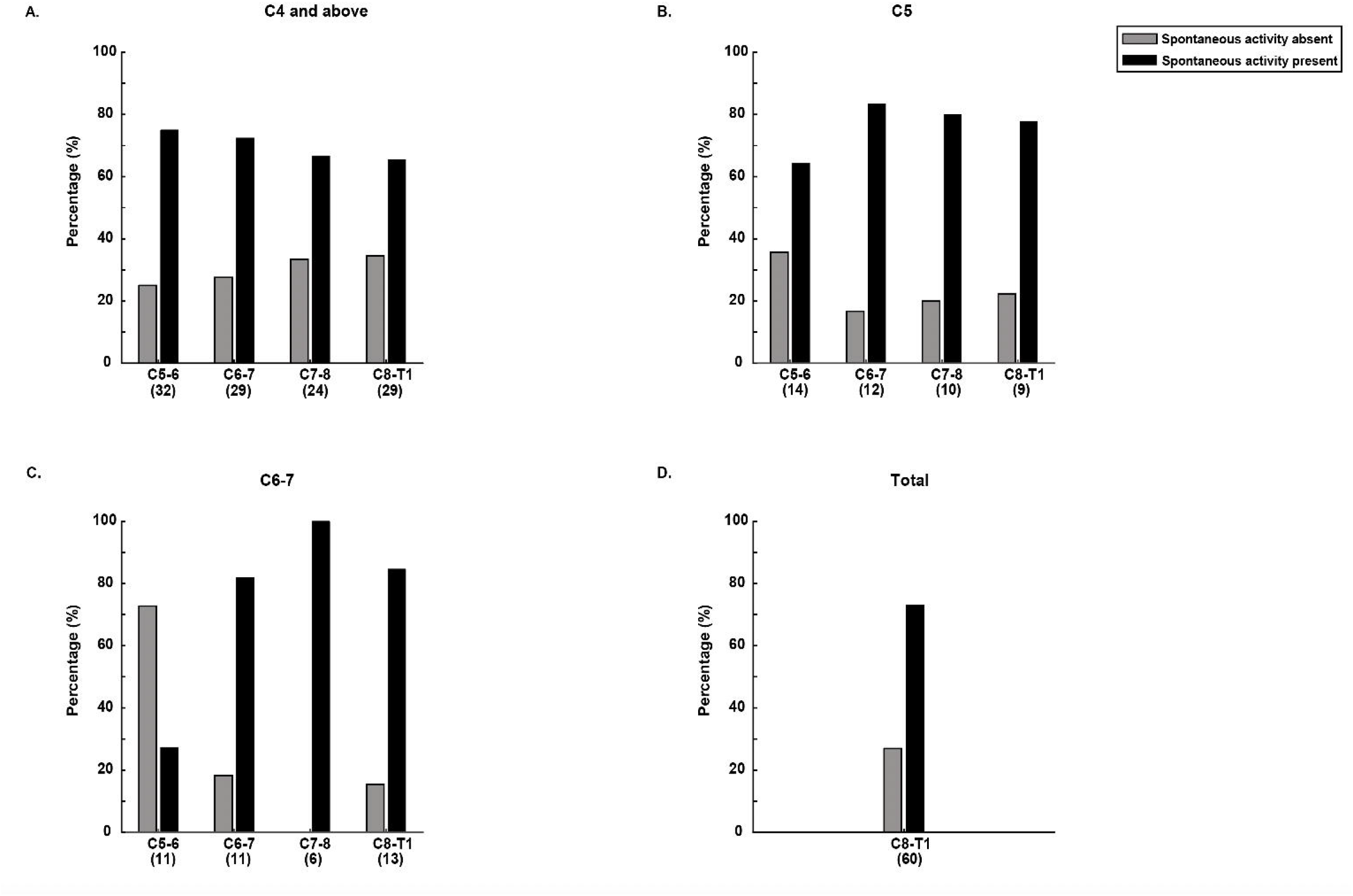
Proportions of denervation potentials observed from segmental spinal levels, stratified by neurological level of injury. Panel D represents the pooled data from the C8-T1 segments. The numbers in parentheses below the segmental levels represent the number of observations recorded, as not all segmental levels were sampled for each limb tested.

The association between needle EMG findings at the C8-T1 segments and median motor CMAP amplitude, is presented in Table 3. Median motor studies were performed in 31 limbs, with corresponding evaluation of the C8-T1 segments with needle EMG. Using McNemar’s test, no significant difference was observed (p=0.181), meaning that EMG abnormality was not associated with a significant probability of also observing abnormal CMAP amplitude, with 14 discordant pairs observed.

**Table 3.** Contingency table of matched pairs for spontaneous activity on needle EMG and CMAP amplitude (n=31).

| **Median CMAP-APB** |  |  |  |
| --- | --- | --- | --- |
| **EMG of C8-T1 myotomes** |  | *Normal* | *Abnormal* |
|  | *Normal* | 8 | 4 |
|  | *Abnormal* | 10 | 9 |

EMG: electromyography; CMAP: compound muscle action potential; APB, abductor pollicis brevis.

The frequency of denervation potentials at contiguous segments caudal to the clinical neurological level of injury is presented in Figure 2. As can be observed, the majority of subjects demonstrated evidence of multi-segmental LMN involvement, including in spinal segments distant from the clinical lesion epicentre. Additionally, 23.1% of participants demonstrated evidence of LMN involvement at the segment proximal to the clinical lesion epicentre.

**Figure 2.**
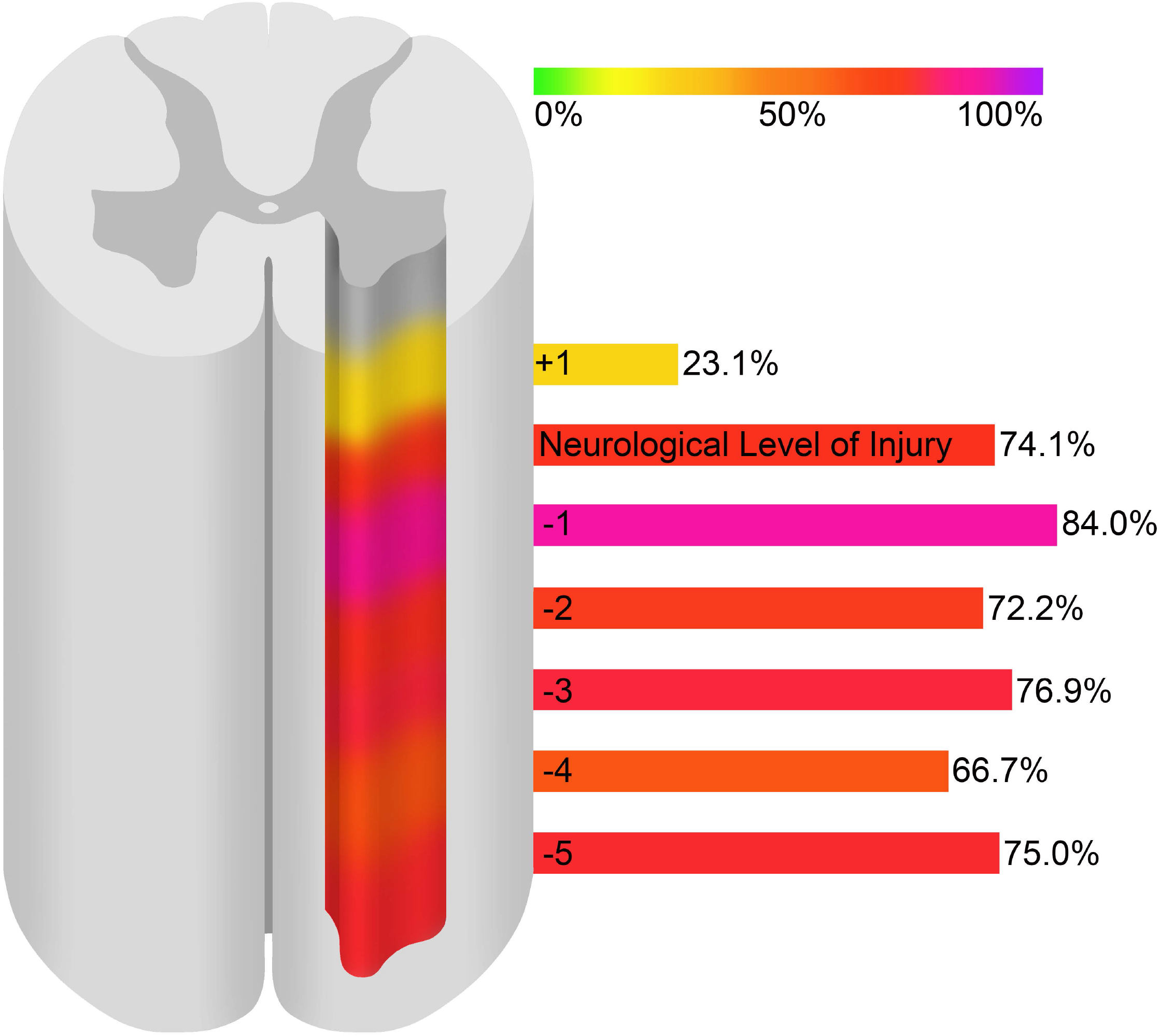
Schematic representation of the proportion of individuals who demonstrated evidence of denervation potential in spinal segments distant from the clinical neurological level of injury, defined by the International Standards for Neurological Classification of Spinal Cord Injury. In some individuals with high tetraplegia, denervation potentials were observed in up to five spinal segments below the clinically defined lesion epicentre.

## Discussion

In this retrospective cohort analysis, we demonstrated a high prevalence of LMN abnormality in spinal segments below the neurological level of injury in individuals early after cervical SCI, including in 74.6% of limbs at the functionally critical C8-T1 myotomes. These results underscore the importance of early of evaluation of the infralesional LMN following cervical SCI, so that interventions such as nerve transfer surgery, are offered and implemented before irreversible muscle fibrosis and atrophy occurs. These findings have implications to the clinical evaluation of the upper limb following cervical SCI, as well as adding to our understanding of the origins of motor dysfunction.

The high frequency of infralesional denervation potentials observed in this study, is similar to our recent analysis of a cohort of individuals with chronic injuries > 2 years duration.^8^ In this previous analysis, we found evidence of LMN abnormality in 87% of muscle groups that are typical recipients for nerve transfer surgery. The present study has perhaps more significant clinical ramifications as this cohort of individuals was evaluated in the sub-acute period after injury (mean 3.3 months). As LMN denervation results in irreversible motor endplate changes within 12-18 months,^21,22^ our data support that individuals should be evaluated with electrodiagnostic testing early after injury, to avoid a scenario whereby they might lose the opportunity for time sensitive intervention. An analogous clinical scenario is brachial plexus injury. Individuals with brachial plexus injury are typically screened with electrodiagnostic studies before 6-months following injury. If complete denervation is observed in a muscle group, then reconstructive surgery may be offered to avoid the irreversible effects of chronic denervation.^18^

The timing of nerve transfer surgery intervention presents a decision-making conundrum for clinicians and individuals with SCI. Those who have no evidence of infralesional LMN abnormality in muscle groups that are the targets of nerve transfer surgery can theoretically delay decision-making regarding intervention indefinitely, with the understanding that infralesional LMNs will remain “preserved”.^5^ Delaying surgery allows more time for spontaneous neurological recovery, which may still occur one year after injury, although this is more likely in individuals with motor incomplete injuries and likelihood of achieving functional strength in clinically relevant upper limb muscles has been reported to be very low (3% of individuals regained > 3 finger flexion strength between 6-12 months after injury, if initial strength grade was 0-2).^23,24^ In the present study, infralesional LMN abnormality was observed in the majority of limbs tested, especially in the functionally critical C8-T1 myotomes, which are responsible for grasp function. This means that individuals who opt to wait for spontaneous recovery may lose their candidacy or have a sub-optimal response to nerve transfer surgery.^6^

The pathophysiological significance of the observed denervation potentials in this study is not entirely clear. Denervation potentials represent muscle membrane instability and are not indicative of the severity of a neuropathic lesion or whether compensatory collateral sprouting is occurring. Denervation potentials may be representative of complete or partial axonal loss.^25^ Using EMG findings to evaluate motor neuron loss is particularly challenging in SCI, as muscles are typically not under volitional control due to processes that are mediated centrally (e.g. damage to the corticospinal tracts). In conventional clinical EMG, assessment of spontaneous activity is combined with qualitative and semi-quantitative assessment of motor unit potential morphology, firing rates and interference patterns, which allow for inferences to be made about the severity and chronicity of LMN lesions.^26^ Size and stability parameters of motor unit potentials are also helpful in determining the degree to which collateral sprouting is taking place.^25^ In the absence of volitional control, only generalizations about whether a muscle is normal or abnormal can be made. The severity and chronicity of LMN lesions cannot be determined on the basis of this data.

Whether the denervation potentials observed reflect direct injury to LMNs or processes related to trans-synaptic degeneration is also a matter for debate. Trans-synaptic degeneration, coined by McComas decades ago,^27^ is a term for the slow degradation of LMNs over time due loss of supraspinal input from an upper motor neuron lesion. In SCI, trans-synaptic degeneration has been attributed to reduced motor neuron numbers, dysregulation of the neuromuscular junction and muscle fibre/energetic changes that have been observed in both animal and human models.^28-33^ Whether the denervation potentials observed in our study are reflective of a direct injury to the anterior horn cells, or trans-synaptic degeneration is not readily determinable. The time course of trans-synaptic degeneration is unknown, but individuals in our study were evaluated relatively early after their injuries. In their large cohort study of individuals with cervical SCI, Van De Meent et al.,^34^ documented recovery profiles of the ulnar CMAP amplitude at serial intervals after the first year of SCI (representing the C8-T1 segments). They showed that the CMAP amplitude drops almost immediately after SCI, but then recovered to above baseline at 12 months post-injury. They speculated that trans-synaptic degeneration, followed by compensatory collateral axonal sprouting, was responsible for decrease and subsequent increase in CMAP amplitude. On examination of this data, the mean baseline CMAP within a month of injury in their study was 2.7-3.7 mV for individuals that were initially classified as motor complete injuries. As lower limit of normal ADM CMAP amplitude is generally accepted to be 6.0 mV,^20^ this suggests that a large proportion of individuals experienced a direct LMN injury at C8-T1 spinal segments, rather than experiencing gradual trans-synaptic-degeneration. These data, in addition to our observations suggest that in many individuals with cervical SCI, there may not be an “intact” infralesional zone with preservation of the LMN.

In support of a multi-segmental longitudinal spinal injury zone, our data from Figure 2 demonstrated the majority of participants have a LMN injury zone that is several segments caudal to the clinically defined neurological level of injury. In some cases, for those with high tetraplegia (C4 and above) the LMN injury zone encompassed spinal segments contiguously to C8-T1. We previously proposed that spinal cord imaging may be a useful adjunct to determine the longitudinal extent of the lesion, as it relates to segmental anterior horn cell damage.^35^ Imaging studies of spinal cord have demonstrated that the longitudinal extent of the spinal lesion extends over multiple spinal segments and that the longitudinal length of edema is negatively associated with clinical outcomes, ^36,37^ although no study to date has directly examined the relationship between the longitudinal extent of a cervical lesion and anterior horn cell involvement.

The other zone of injury that is of immediate clinical relevance is the supralesional zone, specifically the spinal segment that is just rostral to the lesion epicenter, which is the basis for determining the neurological level of injury, according to ISNCSCI. At this level, the muscles supplied may still demonstrate normal or functionally useful strength (manual muscle strength ≥ 3) but may have LMN abnormality. In support of this, our data demonstrated that in those with injuries at C5 and C6/7, the lesion epicentre (C5-6 level) was 64.3% and 25.0% respectively, despite having manual muscle strength ≥ 3 at that level. This has implications to donor nerve selection in nerve transfer surgery. Potential donors for the common reconstructive options are typically derived from the C5-6 segmental levels.^38^ Mandeville et al.^39^ showed that an estimate of motor unit number derived from the needle EMG interference pattern of a donor muscle, was predictive of recipient muscle strength one year after nerve transfer surgery. It has long been postulated that the health of the donor nerve is paramount in supplying a viable pool of axons for recipient muscle reinnervation. Our data suggest that the electrophysiological health of potential donor muscles may be compromised in a significant number of individuals, which may limit candidacy for nerve transfer surgery.

Beyond nerve transfer surgery, infralesional LMN abnormality may be implicated more broadly in predicting functional status after SCI. Hupp et al.^40^ demonstrated that electrophysiological parameters, including NCS responses, can be included in models to enhance functional outcome predictability, using the Spinal Cord Independence Measure as a dependent variable. To our knowledge, no study to date has directly examined the influence of LMN health on outcomes that are specific to the upper limb. We speculate that LMN dysfunction could be one explanation for the difficulty in predicting upper limb motor prognosis, with only 45% of individuals achieving grade > 2 muscle strength at 12 months, in the spinal segment directly caudal to the injury level, when initial strength is grade 0.^41^ It is possible, that infralesional LMN dysfunction could be predictive of the variance in upper limb measures such as the Upper Extremity Motor Score or functional batteries. Based on the present results, it is logical that future studies should directly evaluate that relationship between LMN denervation and upper limb motor outcome.

### Limitations

As mentioned in sections above, the main limitation of the data presented is the ordinal nature and non-specificity of the observed denervation potentials. We provided justification in our Methods section regarding our selection of EMG parameters over more quantitative CMAP parameters, to reflect LMN abnormality. However, others have suggested that the CMAP amplitude may provide a more robust measure of LMN health. Jain et al.^7^ demonstrated that CMAP amplitude, rather than the presence of denervation potentials, was more predictive of a positive response of recipient muscle groups to intra-operative stimulation. Dibble et al.^6^ recently retrospectively reviewed 52 nerve transfers cases and showed that a combination of normal CMAP and EMG findings in recipient muscles, were associated with the greatest motor recovery. However, the CMAP amplitude is problematic as a biomarker for both physiological and technical reasons. The CMAP amplitude is a gross measure of the summed electrical activity of muscle fibre action potentials and may remain normal until up to 50% of the motor neuron pool is lost, due to collateral sprouting.^42^ Additionally, CMAPs derived from clinically relevant donor and recipient muscles are not widely used and have limited reliability and normative data. Finally, CMAPs that are available (e.g. median and ulnar motor studies) can only be used as proxies for muscles that are directly of interest (anterior and posterior interosseous innervated muscles). In our data, we did not find a statistically significant probability that the abnormal EMG data at C8-T1 were associated with an abnormal CMAP (Table 3). Therefore, using the median CMAP as a proxy for other clinically relevant C8-T1 muscle groups may not be entirely valid. Ultimately, more robust quantitative measures of LMN health will be required, due to the limitations of both conventional clinical EMG and NCS, in this clinical scenario. Future studies could evaluate the utility of neurophysiological techniques such as motor unit number estimation, which have been previously used in SCI.^43^

## Conclusion

In this retrospective analysis of LMN abnormality in SCI, we demonstrated a high frequency of denervation potentials involving spinal segments caudal to the lesion epicenter. Additionally, we demonstrated evidence of denervation potentials in segments rostral to the lesion epicenter in many individuals. These findings likely have implications to the candidacy, timing and success of nerve transfer surgery in SCI. As described, these findings may also have broader implications to motor prognostication and functional outcomes after SCI. The limitations of the data as qualitative marker of LMN abnormality have been described, providing justification for more robust and quantitative neurophysiological LMN biomarkers in future studies.

## Data Availability

All data produced in the present study are available upon reasonable request to the authors.

## Acknowledgements

MJB would like to acknowledge Mr. Emmanuel Ogalo for helping design Figure 1. AAA would like to acknowledge support from the Foundation for Physical Medicine and Rehabilitation (Richard S. Materson ERF New Investigator Research Grant). CKF would like to acknowledge support from the Belle Carnell Regenerative Neurorehabilitation fund.

## Authorship confirmation/contribution statement

Lead author MJB contributed to conceptualization, methodology, formal analysis, writing and funding acquisition. Authors AAA, CD, AKH, CM, RO, RS, SS and EB contributed to conceptualization, review/editing and data collection. Author HW contributed to data collection and analysis. Author CKF contributed to conceptualization, reviewing/editing and data collection.

